# Social Distancing Interventions in the United States: An Exploratory Investigation of Determinants and Impacts

**DOI:** 10.1101/2020.05.29.20117259

**Authors:** Shenyang Guo, Ruopeng An, Timothy D. McBride, Danlin Yu, Linyun Fu, Yuanyuan Yang

**Affiliations:** Brown School, Washington University in St. Louis; Department of Earth & Environmental Studies, Montclair State University, 1 Normal Ave., Montclair, NJ 07043, USA

**Author notes:** Corresponding Author, Campus Box 1196, One Brookings Drive, St. Louis, MO 63130, USA.

## Abstract

**Background:** To combat the Covid-19 pandemic in the United States, many states and Washington DC enacted Stay-at-Home order and nonpharmaceutical mitigation interventions. This study examined the determinants of the timing to implement an intervention. Through an impact analysis, the study explored the effects of the interventions and the potential risks of removing them under the context of reopening the economy.

**Method:** A content analysis identified nine types of mitigation interventions and the timing at which states enacted these strategies. A proportional hazard model, a multiple-event survival model, and a random-effect spatial error panel model in conjunction with a robust method analyzing zero-inflated and skewed outcomes were employed in the data analysis.

**Findings:** To our knowledge, we provided in this article the first explicit analysis of the timing, determinants, and impacts of mitigation interventions for all states and Washington DC in the United States during the first five weeks of the pandemic. Unlike other studies that evaluate the Stay-at-Home order by using simulated data, the current study employed the real data of various case counts of Covid-19. The study obtained two meritorious findings: (1) states with a higher prevalence of Covid-19 cases per 10,000 population reacted more slowly to the outbreak, suggesting that some states may have missed the optimal timing to prevent the wide spread of the Covid-19 disease; and (2) of nine mitigation measures, three (non-essential business closure, large-gathering bans, and restaurant/bar limitations) showed positive impacts on reducing cumulative cases, new cases, and death rates across states.

**Interpretation:** The opposite direction of the prevalence rate on the timing of issuing the mitigation interventions partially explains why the Covid-19 caseload in the U.S. remains high. A swift implementation of social distancing is crucial— the key is not whether such measures should be taken but when. Because there is no preventive vaccine and because there are few potentially effective treatments, recent reductions in new cases and deaths must be due, in large part, to the social interventions delivered by states. The study suggests that the policy of reopening economy needs to be implemented carefully.

---

As of May 1, the total confirmed cases of Covid-19 reached 3.4 million worldwide, causing nearly 240,000 deaths; the U.S. experienced the highest disease burden, with over 1.1 million confirmed cases (i.e., 33.5% of the world total) and more than 65,000 deaths (i.e., 27.8% of the world total).^1,2^ On March 13, 2020, the President of United States declared a National Emergency to combat the coronavirus pandemic. Following his declaration, 42 States and Washington DC enacted Stay-at-Home orders, with California starting the earliest (3 AM 3/19/2020, U.S. Eastern Time) and South Carolina starting the latest (5 PM on 4/7/2020, U.S. Eastern Time). These orders covered approximately 94.7% of the U.S. population. Closely related to the Stay-at-Home order are various nonpharmaceutical containment interventions aiming to promote social distancing.

Social distancing is an essential component of a public health response to infectious-disease outbreak. A systematic review of influenza pandemics^3^ found that by isolating ill persons, contact tracing, quarantining exposed persons, school closures, workplace closures, and avoiding crowding, distancing helped reduce transmission, delay transmission, and reduce epidemic peak.

Covid-19 is a novel coronavirus disease. Compared to influenza and severe acute respiratory syndrome (SARS), Covid-19 has a more extended incubation period of 5–6 days, though its doubling time (i.e., the speed of the initial spread of the epidemic) or the related serial interval (i.e., the mean time it takes for an infected person to pass on the infection to others) is similar to SARS, which is estimated at 4.4–7.5 days.^4^ A study in China found that among cases known to have traveled from Wuhan before January 23, the time from symptom onset to confirmation was 6.5 days (SD = 4.2 days), and for those who traveled from Wuhan after January 23 when more active surveillance was in operation, the interval was reduced to 4.8 days (SD = 3.03 days).^5^ These data suggest that, to prevent the spread of Covid-19, policymakers must implement mitigation strategies – if they choose to do so – in a speedy fashion. In the absence of speedy implementation, the prevalence is likely to increase rapidly. Therefore, the timing of enacting mitigation interventions plays a vital role in public health. Given that patients during the incubation period are asymptomatic, responding to confirmed cases quickly has the potential to make a significant difference in subsequent disease levels.

The purpose of this study is to undertake a correlational analysis of the determinants and impacts of U.S. mitigation measures used in the first five weeks of the pandemic. The study intends to address two research questions. Using an early period in the pandemic (i.e., from 3/13/2020 to 4/7/2020), one question focuses on the determinants of the implementation timing of mitigation strategies. Why did some states in the United States respond rapidly to the pandemic while others did not? Of various determinants, we are primarily concerned with the prevalence rate of the confirmed cases vis-à-vis the time when each mitigation strategy was enacted. Delay in taking intervention actions during this period may directly affect the speed and scope of the Covid-19 spread.

The second question focuses on the correlations between the timing of enacting mitigation and the serial incidence of Covid-19 cases and deaths. For this preliminary assessment of the impact of mitigation interventions, the observational window was extended to 4/15/2020. The impact study aims to address a key question: Is an initial response observed? Precisely during this period, many states had begun to reopen their economies and public life. By May 11, 32 states chose to do so.^6^ In light of these decisions, the purpose of this paper is to explore the initial impact of nonpharmaceutical mitigation interventions in the event that re-opening the economy is associated with a second wave of Covid-19 disease and a second round of mitigation becomes necessary.

## Methods

### Study Window

In this study, we define the enactment of social distancing strategies as the exact hour at which a mitigation intervention was implemented, as stipulated by state government documents. The study window is defined as the period from the time when a National Emergency was declared to the time when the last enacting state issued a Stay-at-Home order. More precisely, the zero hour for this study is 12 PM March 13, U.S. Eastern Time, and the ending hour is 6 PM April 7, which is one hour after the last enacting state (i.e., South Carolina) implemented its Stay-at-Home order (5 PM April 7, U.S. Eastern Time). States declined to issue a mitigation order are considered censored at 6 PM on April 7, 2020. Within this 25.25-day or 606-hour window, we study not only whether a state enacted mitigation interventions, but also the exact times at which these interventions were implemented. The timing of the National Emergency is a natural starting point for the study because, before that date, none of the migration strategies was enacted in the United States. A special caveat should be put on the word “zero”: it does not mean that the Covid-19 pandemic started at that point, because as Anderson et al. pointed out: “Most countries are likely to have spread of COVID-19, at least in the early stages, before any mitigation measures have an impact.”^6^

### Data Source

Qualitative content analysis was used to analyze large quantities of text data from governmental documents. We examined all relevant documents (a total of 1,470 executive orders) pertinent to the social distancing orders from the state-government websites for all 50 states and Washington DC. A content-coding/analysis^7^ was performed by two researchers. Review and coding guidelines were developed in advance. The two reviewers separately examined all documents, and resolved discrepancies in coding by consensus in a discussion. Considering variation across states in the frequency of use of alternative social distancing strategies and prior literature on social distancing, we extracted information on 9 types of the mitigation interventions: Stay-at-Home order, strengthened Stay-at-Home order, public school closure, all school closure, large-gathering ban of more than 10 people, any gathering ban, restaurant/bar limit to dining out only, non-essential business closure, and mandatory self-quarantine of travelers. The exact timing to implement each of these 9 nonpharmaceutical interventions (i.e., the hour when a specific strategy was implemented) was recorded. A state that did not enact a particular intervention by the end of the study window (i.e., 6 PM April 7) was considered censored for that strategy and was coded as having the longest time of 606 hours on length of time it took to enact the measure. Censoring under the current setting refers to the fact that the event of interest did not occur by the end of the study window but might occur in future.

The theoretical model guiding the selection of potential determinants for inclusion in our models of social distancing orders is the “social determinants of health” (SDOH) framework.^8^ Originally developed by WHO, the framework emphasizes the strong relationship between societal factors and health outcomes, especially crucial activities needed to close gaps and inequalities in health care concerning socioeconomic status, gender, tradition/culture, and race/ethnicity. To adapt this framework for the U.S., the National Academies of Science, Engineering, and Medicine organized a special group to further articulate strategies for integrating social care into the delivery of health care to improve the nation’s health. Report of the Academies was published on September 25, 2019. Immediately following its publication, a Congressional Briefing was convened to discuss the implementation of SDOH-related strategies.^9^ The Academies’ report underscores five key health care activities to better address the SDOH and integrate social care with health care: awareness, adjustment, assistance, alignment, and advocacy. These activities help us to identify important variables for the determinant study.

Under the guidance of the five key SDOH activities and based on the available census data and other published statistics, we identified the following five blocks of state-level variables for inclusion as potential determinants in modeling social distance orders: demographic characteristics, economic well-being information, public heath infrastructure, information related to politics, and international connectivity. To measure state governmental awareness of the Covid-19, we included in the study a time-varying covariate of cumulative Covid-19 cases per 10,000 population one-day before the time when a mitigation strategy was enacted. This measure is analogous to the prevalence rate of Covid-19 disease.

To explore the impacts of mitigation interventions, we conducted statistical analyses that link the timing of each mitigation order to various case counts of Covid-19. This is a correlational rather than causal analysis, due to lacking an accurate or even a proxy estimate of counterfactual. A counterfactual in the current setting refers to “What would have happened had we not enacted a mitigation strategy?” The outcome data come from Johns Hopkins University Coronavirus Data Stream that combines WHO and CDC case data.^10^ Based on the original input data, we employed 9 variables in the investigation: cumulative cases, cumulative deaths, new cases, new deaths, cumulative cases per 10,000 population, cumulative deaths per 10,000 population, cumulative new cases per 10,000 population, cumulative new deaths per 10,000 population, and death rate defined as the number of cumulative deaths divided by the number of cumulative cases. These measures are computed as of the time of this study and may underestimate the actual cases and deaths. The study employed daily counts on each of the 9 outcome measures from March 11 (two days before the National Emergency was declared) to April 15 (about one week after the last state enacted the Stay-at-Home order).

### Survival Analysis and the Multiple-Event Survival Model

The analysis of determinants of each mitigation strategy employed a proportional hazards model.^11, 12^

The analysis of overall determinants pulled together all nine episodes of mitigation interventions and employed the WLW model. This model is designated by the first initials of the developers’ last name.^13,14^

The WLW model is a robust approach developed to control for the autocorrelations of study events. Under the framework of Cox proportional hazards model, the hazard function for the *i*^th^ state to enact the *k*^th^ type of mitigation intervention is expressed as

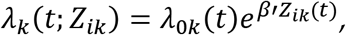

where *Z_ik_* = (*Z*_1*ik*_,…, *Z*_*pik*_)′ denotes the covariate vector for the *i*^th^ state with respect to the *k*^th^ type of order, λ_0k_(*t*) with (*k* = 1,…, *K*) is the unspecified baseline hazard function, *K* is the maximum number of order types in a state, and *β* = (*β*_1_,…, *β_p_*)′ is a p × 1 vector of unknown regression parameters.

Let *T_ik_* be the time when the *k*^th^ type of event occurs in the *i*^th^ state, and let *C_ik_* be the corresponding censoring time. Define *X_ik_* = min (*T_ik_, C_ik_*), and *Δ_ik_* = I(*T_ik_<C_ik_*), and *I_(A)_* indicates, by the values one versus zero, whether or not the event A occurs. When the observations within the same state are independent, the partial likelihood function for *β* is

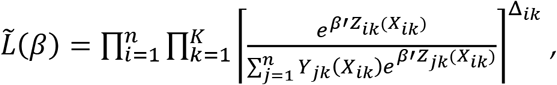

where *Y_ik_*(*t*) = *I*(*X_ik_* ≥ *t*). The corresponding score function is

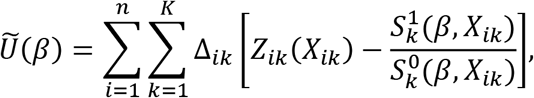

where 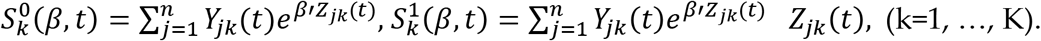. By this specification, we obtain the unique estimator 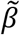 by solving [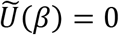].

When the observations within the same state are not independent, the derivative matrix 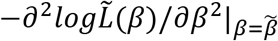 does not provide a valid variance-covariance estimator for *Ũ*(*β*). The WLW method approximates *Ũ*(*β*) with a sum of *n* independent and identically distributed random vectors. By doing this, the model establishes the asymptotic normality of *Ũ*(*β*) and obtains its limiting covariance matrix. The asymptotic distribution for 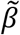 follows from the Taylor series expansion.

Under the WLW specification, for large *n* and relatively small *K*, the statistic *Ũ*(*β*) is approximately *p*-variate normal with mean zero and estimated covariance matrix

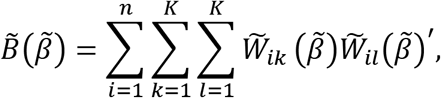

where

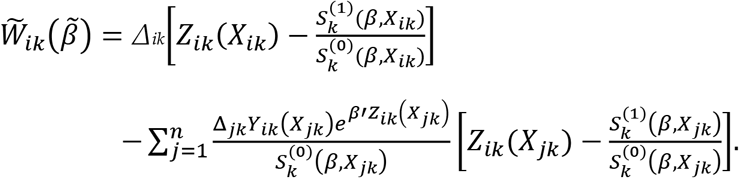

Furthermore, the estimator 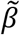 is approximately *p*-variate normal with mean β and estimated covariance matrix 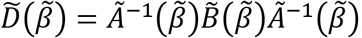, where

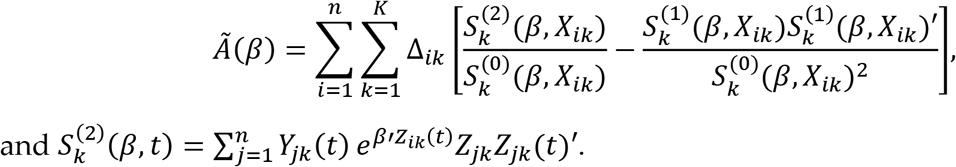

and 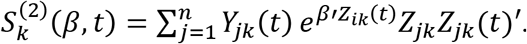.

### The Inverse-Normal Transformation (INT) and the Indirect INT Method

To analyze zero-inflatd and skewed outcome variables (i.e., all Covid-19 caseload data), we employed the inverse-normal transformation approach. This approach is frequently practiced in genome-wide association studies in which skewed residuals are common and, hence, violate the normality assumption embedded in regression analysis. Suppose *u* has a skewed distribution. Let rank(*u_i_*) denote the sample rank of *u_i_* when the measurements are placed in ascending order. The rank-based inverse normal transformation (INT) is defined as:

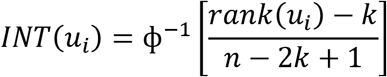

Here ф^−1^ is a normal density function, *k* ∈ (0, ½) is an adjustable offset, and *n* is the sample size. By default, the Blom offset of *k* = 3/8 is adopted.

The impact study employed a newly developed method called indirect-INT that shows efficient and unbiased properties in Genome-Wide Association Studies.^15^ Denoting *y_it_* the outcome variable *y* for the *i^t^*^h^ state at time *t*, *t* ∈ {1,…, *T*}, the analysis followed the steps described below: (1) Separately for each time point t ∈ {1,…, 𝑇}, regress each *y_it_* on the time-invariant covariates *w_i_* to obtain the residuals *ε_it_*; (2) Conduct INT on the residuals *Z_it_* ≡ *INT*(*ε_it_*) to obtain the Z-scores, again separately for each time point *t*; (3) Regress each mitigation strategy on the time-invariant covariates *w_i_* to obtain the residuals; and (4) Fit the full random-effect spatial error panel model for the *Z_it_*.

### Method of the Impact Analysis Controlling for Spatial Autocorrelations

Residuals of a regression model with spatial data often show spatial autocorrelation that is caused by either the spatial autocorrelated dependent variables (endogenous interaction) or omitted spatially autocorrelated predictors (error interaction).^16, 17^ The predictors might also be spatially autocorrelated (exogenous interaction), but it will not cause residual spatial autocorrelation. To address the autocorrelation, we employed a spatial panel model. A full version of the model can be expressed as:

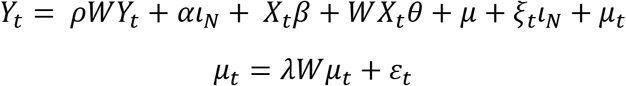

where *Y_t_* and *X_t_* are the dependent and independent variables at time *t*, respectively. *β* is the vector of coefficients. *WY_t_, Wµ_t_* and *WX_t_* are the three interactions. *ρ, θ* and *λ* are their coefficients. *W* is a spatial configuration matrix and is defined using a sphere of influence (SOI) approach^18^ that defines for the *i*^th^ unit, let *r_i_* be the distance from *i* to its nearest neighbor, and *C_i_* the circle centered on *i* with the radius of *r_i_. i* and *j* are neighbors when *C_i_* and *C_j_* intersect. *µ_t_* is the error term. *ιN* is a vector of value ones. *µ* and *ξ_t_* are individual and time-specific effects. *ε_t_* is a well-behaved random noise.

Because the sources of residuals’ spatial autocorrelation can only be weakly identified^17^, modeling either the endogenous (spatial lag) or error interaction (spatial error) would suffice. In this study, we are interested in how the mitigation strategies impact the spread of covid-19. There are likely omitted spatially autocorrelated predictors. Therefore, we employed a random-effect spatial error panel model in conjunction with an indirect-INT in the final analysis.

## Results

### Factors Associated with the Implementation of Nonpharmaceutical Mitigation Interventions

Figure 1 presents the descriptive information about the implementation of these mitigation interventions for all 50 states and Washington DC. The most important strategy is the Stay-at-Home order, which was enacted by 42 states and Washington DC. The earliest state to implement this was California. Most states issued a Stay-at-Home order between March 23 and March 28. However, eight states never chose to implement the order. The map shows the geographic distribution of timing categories, with lighter colors indicating earlier enactments of Stay-at-Home. The most frequently invoked intervention was public school closure, which was enacted by all 50 states and Washington DC. Most states issued a school closure order between March 13 to 18. The second most frequently invoked intervention was restaurant/bar closure or limitation, which was issued by 49 states and Washington DC. A strengthened order refers to mitigation more restrictive than Stay-at-Home, including the enforcement of Stay-at-Home and grocery shopping limit. Only five states issued a strengthened order. Because of the comparatively low use of strengthened order, we did not include it in Figure 1. Of these five states, Hawaii implemented the strengthened order on 6 AM April 1, U.S. Eastern Time, with a length of time of 450 hours, and the remaining four states (Indiana, Massachusetts, New York, and Ohio) implemented strengthened orders on 12 AM April 7, U.S. Eastern Time, with a length of time of 588 hours.

**Figure 1:**
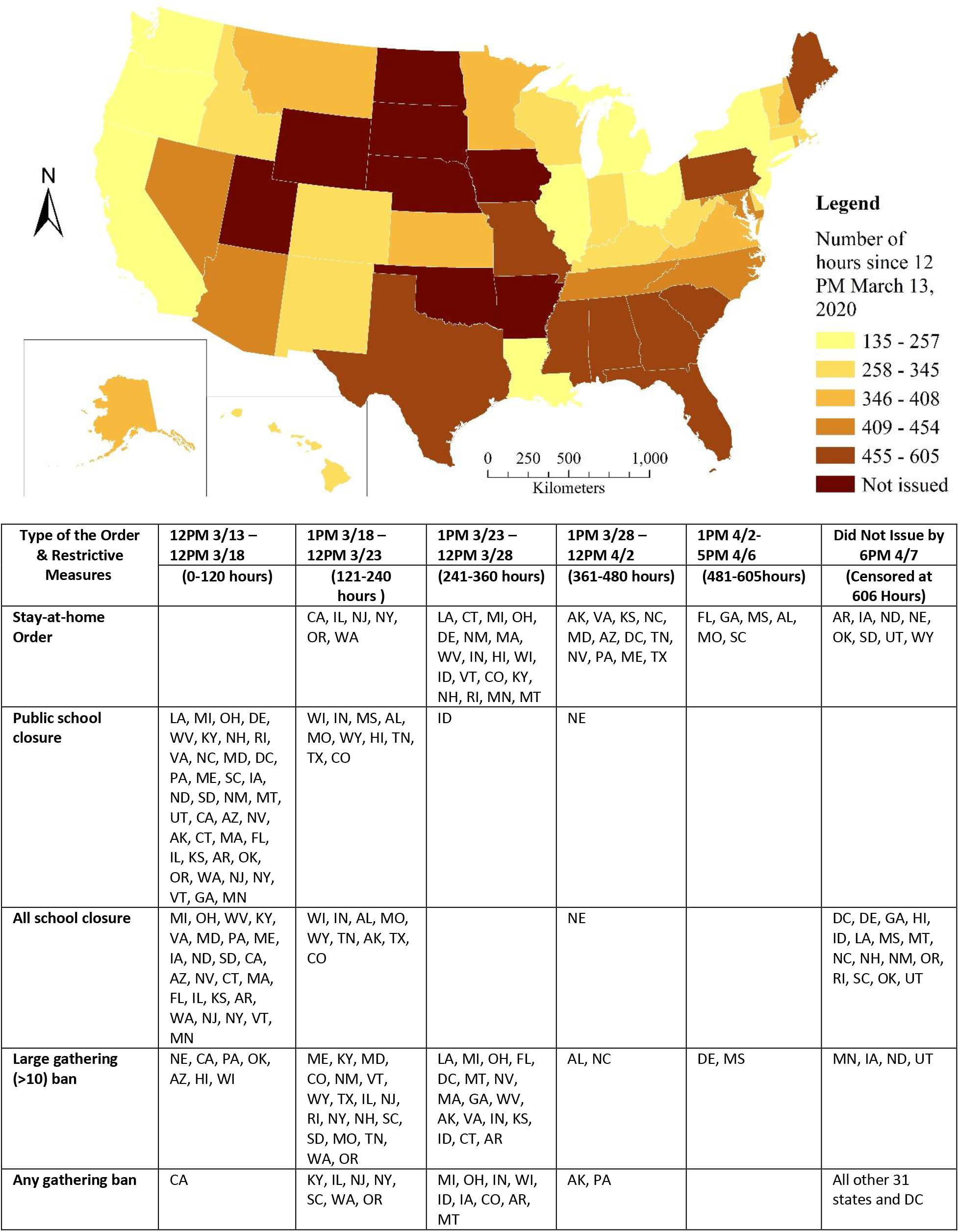
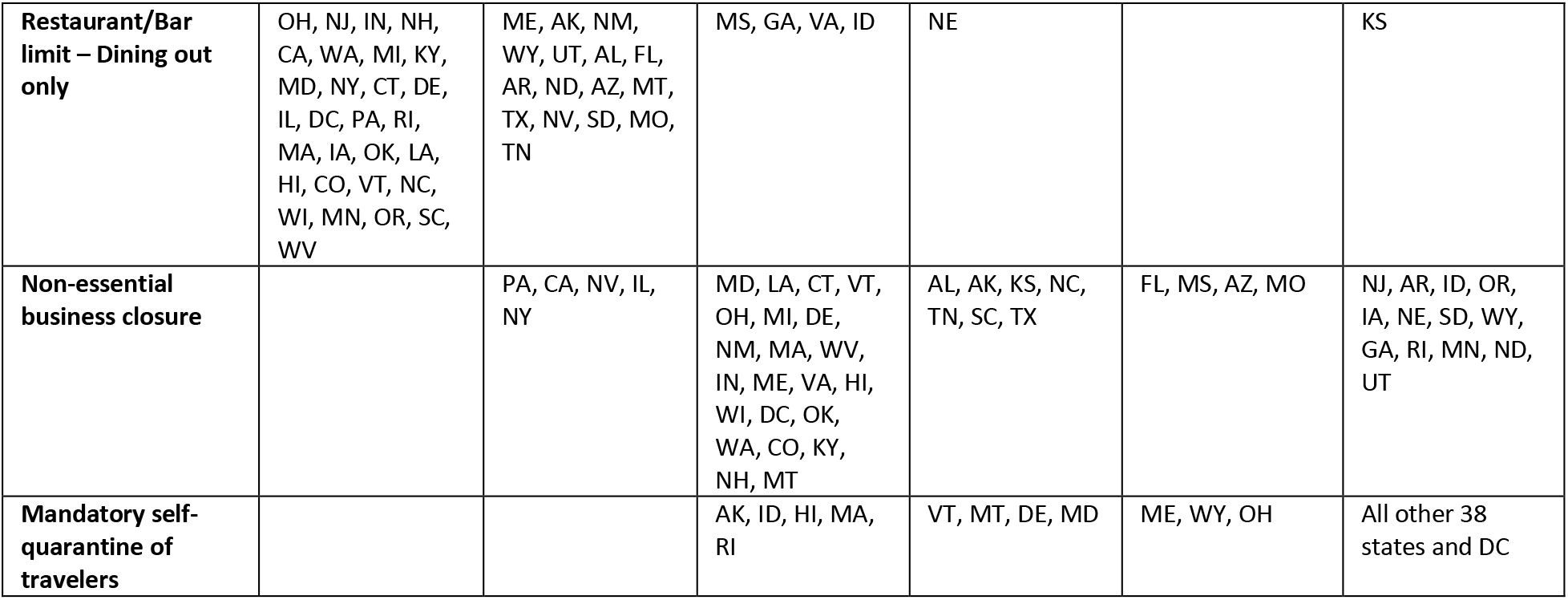
Timing Enacting the Stay-at-Home Order and Restrictive Measures

The results of the estimated models for each of the nine mitigation interventions are presented in Table 1. Of nine models, the strengthened-order model did not fit the data well, as the model chi-square was 12.66 (p = .2434). A poor fit of the strengthened-order model was expected, because only five states, out of 51, invoked this order. The results of the remaining eight models are summarized below.

**Table 1.** Proportional Hazard Models Studying Determinants of the Timing Issuing Each Order & Restrictive Measure

| Variable | Descriptives | Stay-at-Home Order |  |  | Strengthened Order |  |  | Public School Closure |  |  | All School Closure |  |  |
| --- | --- | --- | --- | --- | --- | --- | --- | --- | --- | --- | --- | --- | --- |
|  | Mean (SD) | Hazard Ratio |  | 95% CI | Hazard Ratio |  | 95% CI | Hazard Ratio |  | 95% CI | Hazard Ratio |  | 95% CI |
| Covid19 cases per 10,000 population one day prior | 2.14 (1.66)<sup>a</sup> | 0.449 | *** | (.31, .65) | 1.073 | + | (.99, 1.16) | 0.040 | ** | (.00, .32) | 0.023 | ** | (.00, .27) |
| GDP Per Capita 2019 | 64785.29<br>(23714.35) | 1.000 | * | (1.00, 1.00) | 1.000 |  | (.99, 1.00) | 1.000 |  | (.99,1.00) | 1.000 |  | (.99, 1.00) |
| % African American & Hispanic 2018 | 23.94 (14.07) | 0.998 |  | (.95, 1.04) | 0.876 |  | (.67, 1.14) | 0.998 |  | (.95, 1.04) | 1.045 |  | (.97, 1.12) |
| Poverty Rate 2018 | 12.86 (2.78) | 1.253 | + | (.97, 1.62) | 1.502 |  | (.54, 4.17) | 1.096 |  | (.87, 1.38) | 1.008 |  | (.71, 1.43) |
| % Uninsured 2018 | 8.10 (3.09) | 0.787 | ** | (.68, .92) | 0.707 |  | (.39, 1.30) | 0.856 | + | (.73, 1.01) | 0.659 | ** | (.51, 0.85) |
| Unemployment Rate Feb. 2020 | 3.51 (0.83) | 1.541 |  | (.90, 2.63) | 1.005 |  | (.11, 8.82) | 1.419 |  | (.82, 2.44) | 1.607 |  | (.77, 3.37) |
| Hospital Beds per 1000 Pop. 2018 | 2.64 (0.75) | 0.246 | ** | (.09, .68) | 0.313 |  | (.02, 5.95) | 0.897 |  | (.47, 1.72) | 2.109 |  | (.77, 5.77) |
| Public Health Budget Per Capita 2019 | 107.00(71.28) | 0.995 |  | (.99, 1.00) | 0.987 |  | (.95, 1.02) | 0.995 |  | (.99, 1.00) | 0.999 |  | (.99, 1.01) |
| Democratic Governor 2020 (Ref. Republican) | 0.49 (0.50) | 1.741 |  | (.74, 4.08) | 0.143 |  | (.01, 2.85) | 1.230 |  | (.51, 2.96) | 1.321 |  | (.42, 4.12) |
| Number of International Airports | 0.82 (1.14) | 1.267 |  | (.87, 1.84) | 2.729 |  | (.66, 11.22) | 1.016 |  | (.71, 1.45) | 1.240 |  | (.82, 1.88) |
| Likelihood Ratio Chi-squares (df=10) |  | 68.70 *** |  |  | 12.66 (p=.2434) |  |  | 30.10** |  |  | 90.12 *** |  |  |

\*p<.05, \*\*p<.01, \*\*\*p<.001, +p<.1, two-tailed test
Note: a. This is a time-varying variable. Descriptive statistics shown here are only pertinent to the Stay-at-Home order.

Table 1. Continued
| Variable | Large Gathering Ban |  |  | Any Gathering Ban |  |  | Restaurant/Bar Limit |  |  | Non-essential B. Ban |  |  | Mandatory Self-Qua. |  |  |
| --- | --- | --- | --- | --- | --- | --- | --- | --- | --- | --- | --- | --- | --- | --- | --- |
|  | Hazard Ratio |  | 95% CI | Hazard Ratio |  | 95% CI | Hazard Ratio |  | 95% CI | Hazard Ratio |  | 95% CI | Hazard Ratio |  | 95% CI |
| Covid19 cases per 10,000 population one day prior | 0.324 | *** | (.20, .54) | 0.152 | *** | (.06, .38) | 0.052 | ** | (.01, .38) | 0.384 | *** | (.24, .60) | 0.104 | ** | (.02, .49) |
| GDP Per Capita 2019 | 1.000 |  | (.99, 1.00) | 1.000 | ** | (1.00, 1.00) | 1.000 |  | (1.00, 1.00) | 1.000 |  | (.99, 1.00) | 1.000 |  | (.99, 1.00) |
| % African American & Hispanic 2018 | 1.042 | + | (.99, 1.09) | 1.044 |  | (.96, 1.14) | 1.004 |  | (.96, 1.06) | 1.056 | * | (1.00, 1.11) | 0.877 | * | (.77, .99) |
| Poverty Rate 2018 | 0.933 |  | (.74, 1.17) | 1.740 | * | (1.09, 2.78) | 1.081 |  | (.82, 1.42) | 0.958 |  | (.74, 1.23) | 0.790 |  | (.34, 1.83) |
| % Uninsured 2018 | 0.879 | + | (.76, 1.02) | 0.810 |  | (.58, 1.12) | 0.716 | ** | (.59, .87) | 0.791 | ** | (.67, .94) | 0.977 |  | (.65, 1.47) |
| Unemployment Rate Feb. 2020 | 1.627 | * | (.99, 2.65) | 0.991 |  | (.37, 2.67) | 1.372 |  | (.83, 2.28) | 1.610 |  | (.91, 2.84) | 2.621 |  | (.21, 32.41) |
| Hospital Beds per 1000 Pop. 2018 | 0.889 |  | (.43, 1.86) | 1.106 |  | (.23, 5.34) | 0.696 |  | (.34, 1.41) | 0.950 |  | (.41, 2.19) | 0.021 | ** | (.00, .37) |
| Public Health Budget Per Capita 2019 | 0.997 |  | (.99, 1.01) | 0.995 |  | (.98, 1.01) | 0.993 | + | (.99, 1.00) | 0.996 |  | (.99, 1.00) | 1.041 | * | (1.01, 1.08) |
| Democratic Governor 2020 (Ref. Republican) | 0.859 |  | (.43, 1.71) | 4.301 |  | (.71, 26.01) | 0.752 |  | (.36, 1.55) | 0.799 |  | (.35, 1.84) | 0.096 | + | (.01, 1.22) |
| Number of International Airports | 0.969 |  | (.69, 1.36) | 1.279 |  | (.48, 3.38) | 1.244 |  | (.90, 1.72) | 1.175 |  | (.78, 1.76) | 0.064 | * | (.01, .63) |
| Likelihood Ratio Chi-squares (df=10) | 36.33*** |  |  | 63.92*** |  |  | 56.55 *** |  |  | 55.01 *** |  |  | 55*** |  |  |

\*p<.05, \*\*p<.01, \*\*\*p<.001, +p<.1, two-tailed test

#### Determinants of Stay-at-Home order

Covid-19 cases per 10,000 population at the time one day before the enactment – every one-case increase reduces the hazard of enacting the order by 55.1% (p<.001); GDP per capita in 2019 – every $1,000 increase increases the hazard by 3.9% (p<.05); % uninsured in 2018 – every one-percentage-point increase reduces the hazard by 21.3% (p<.01); hospital beds per 1,000 population in 2018—every one-bed increase reduces the hazard by 75.4%.

#### Determinants of public school closure

Covid-19 cases per 10,000 population at the time one day before the enactment – every one-case increase reduces the hazard of enacting the measure of public school closure by 96% (p<.01).

#### Determinants of all school closure

Covid-19 cases per 10,000 population at the time one day before the enactment – every one-case increase reduces the hazard of enacting the measure of all school closure by 97.7% (p<.01); % uninsured in 2018 – every one-percentage-point increase reduces the hazard by 34.1% (p<.01).

#### Determinants of large gathering ban of more than 10 people

Covid-19 cases per 10,000 population at the time one day before the enactment – every one-case increase reduces the hazard of enacting the large gathering ban by 67.6% (p<.001); unemployment rate in February 2020 – every one-percentage point increase increases the hazard by 62.7% (p<.05).

#### Determinants of any gathering ban

Covid-19 cases per 10,000 population at the time one day before the enactment – every one-case increase reduces the hazard of enacting the any gathering ban by 84.8% (p<.001); GDP per capital in 2019 – every $1,000 increase increases the hazard by 14.2% (p<.01); poverty rate in 2018 – every one-percentage-point increase increases the hazard by 74% (p<.05).

#### Determinants of restaurant/bar limit to dining out only

Covid-19 cases per 10,000 population at the time one-day prior to the enactment – every one-case increase reduces the hazard of enacting the measure of restaurant/bar limit by 94.8% (p<.01); % uninsured in 2018 – every one-percentage-point increase reduces the hazard by 28.4% (p<.01).

#### Determinants of non-essential business ban

Covid-19 cases per 10,000 population at the time one day before the enactment – every one-case increase reduces the hazard of enacting the non-essential business ban by 38.4% (p<.001); % of African American and Hispanic population – every one-percentage-point increase increases the hazard by 5.6%; % uninsured in 2018 – every one-percentage-point increase reduces the hazard by 20.9% (p<.01).

#### Determinants of mandatary self-quarantine of travelers

Covid-19 cases per 10,000 population at the time one day before the enactment – every one-case increase reduces the hazard of enacting the mandatary self-quarantine of travelers by 89.6% (p<.01); hospital beds per 1,000 population – every one-bed increase decreases the hazard by 97.9%; public health budget per capita in 2019 – every one-dollar increase increases the hazard by 4.1% (p<.05); the number of international airports – every one-airport increase decreases the hazard by 93.6% (p<.05).

#### Determinants of the overall model

Table 2 presents the results of a multiple-event analysis. Regardless of the type of mitigation, overall, there are three statistically significant determinants of the timing: population in 2019— every 100,000-population increase increases the hazard of enactment by 4% (p<.05), the unemployment rate in Feb. 2020 – every one-percentage-point increase increases the hazard by 50.7% (p<.001), and % of uninsured in 2018 – every one-percentage-point increase reduces the hazard by 11.3% (p<.01).

**Table 2.** The Multiple-event Survival Model Depicting the Overall Determinants of the Timing Issuing All 9 Types of Measures (Stay-at-Home Order, Strengthened Order, Public School Closure, All School Closure, Large Gathering Ban, Any Gathering Ban, Restaurant/Bar Limit, Non-essential Business Ban, and Mandatory Self-Quarantine of Travelers)

| Variable | Descriptives |  | Hazard Ratio | 95% C.I. |
| --- | --- | --- | --- | --- |
|  | Mean | SD |  |  |
| Covid19 cases per 10,000 population one day prior | 3.286 | 7.04 | 0.572 | (.26, 1.25) |
| Population 2019 | 64.361 | 72.96 | 1.004 * | (.99, 1.01) |
| % of Population aged 65 or older | 16.404 | 1.95 | 1.105 + | (.99, 1.24) |
| % African American & Hispanic 2018 | 23.943 | 13.95 | 1.003 | (.98, 1.03) |
| % Asian 2018 | 4.527 | 5.46 | 0.992 | (.97, 1.01) |
| GDP Per Capita 2019 | 64785.290 | 23506.32 | 1.000 | (.99, 1.00) |
| Poverty Rate 2018 | 12.863 | 2.76 | 1.016 | (.90, 1.15) |
| Unemployment Rate Feb. 2020 | 3.514 | 0.82 | 1.507 *** | (1.25, 1.81) |
| Hospital Beds per 1000 Pop. 2018 | 2.635 | 0.74 | 0.813 | (.56, 1.18) |
| % Uninsured 2018 | 8.096 | 3.07 | 0.887 * | (.81, .97) |
| Public Health Budget Per Capita 2019 | 107.000 | 70.66 | 0.998 | (.99, 1.00) |
| Democratic Governor 2020 (Ref. Republican) | 0.490 | 0.50 | 0.927 | (.67, 1.28) |
| % of Votes for Trump in 2016 | 0.488 | 0.12 | 0.133 | (.01, 2.44) |
| Number of International Airports | 0.824 | 1.13 | 0.957 | (.79, 1.16) |
| Likelihood Ratio Chi-squares (df=14) |  |  | 198.78 (p=.0000) |  |

\*p<.05, \*\*P,.01, \*\*\*p<.001, +p<.1, two-tailed test

One of the important findings is the response of states to the Covid-19 prevalence rate. Contradictory to our hypothesis, states with a lower prevalence of Covid-19 cases per 10,000 population reacted more quickly to the outbreak. Some states appear to have been more primed to issue Stay-at-Home orders. They acted while the virus had a low prevalence before it spread widely. This finding is statistically significant in all 8 models. The variable is not significant in the overall model, probably due to the inclusion of other predictors. Given that the doubling time and related serial interval of Covid-19 are short (i.e., 4.4–7.5 days), taking mitigating actions within a short period is crucial. States that experienced less disease responded in a faster fashion. This finding helps explain why new case counts of Covid-19 in the United States remained high for a longer period than other developed economies globally. The finding suggests that even following the National Emergency declaration, some states did not react and missed the opportunity to prevent the spread of Covid-19.

To better understand the effect of prevalence rate on the timing of enacting mitigation interventions, we present the model-based predictions of survival curves for all eight models and descriptive figures in Figure 2. Fig. 2.1 shows the survival functions, which, in the current setting, indicates the proportion of states that did not enact certain measure at a given time. As such, the bottom survival line indicates the fastest speed to enact, while the top survival line indicates the slowest. The figures indicate that States with high prevalence rates enacted the mitigation measures slowly. Fig. 2.2 shows prevalence rates of Covid-19 cases at the time one day before each mitigation intervention was enacted. States on the X-axis are ordered by the timing of the enactment, with the state closer to the origin enacted earlier. As all figures clearly show, states issued later (i.e., those on the right-hand side) had high Covid-19 cases per 10,000 population (i.e., high bars), indicating that the higher the prevalence rate, the slower the speed moving to the enactment. This pattern applies to all mitigation measures.

In summary, the Covid-19 prevalence rate, population size, unemployment, the proportion of uninsured, poverty, level of economic development, proportion of minority people, and public health infrastructure are the key determinants affecting the timing to enact nonpharmaceutical mitigation interventions.

Figure 2.

Model-based Survival Curves and Bar Charts Showing the Impact of Prevalence Rate on Timing of Mitigations

**Fig. 2.1.**
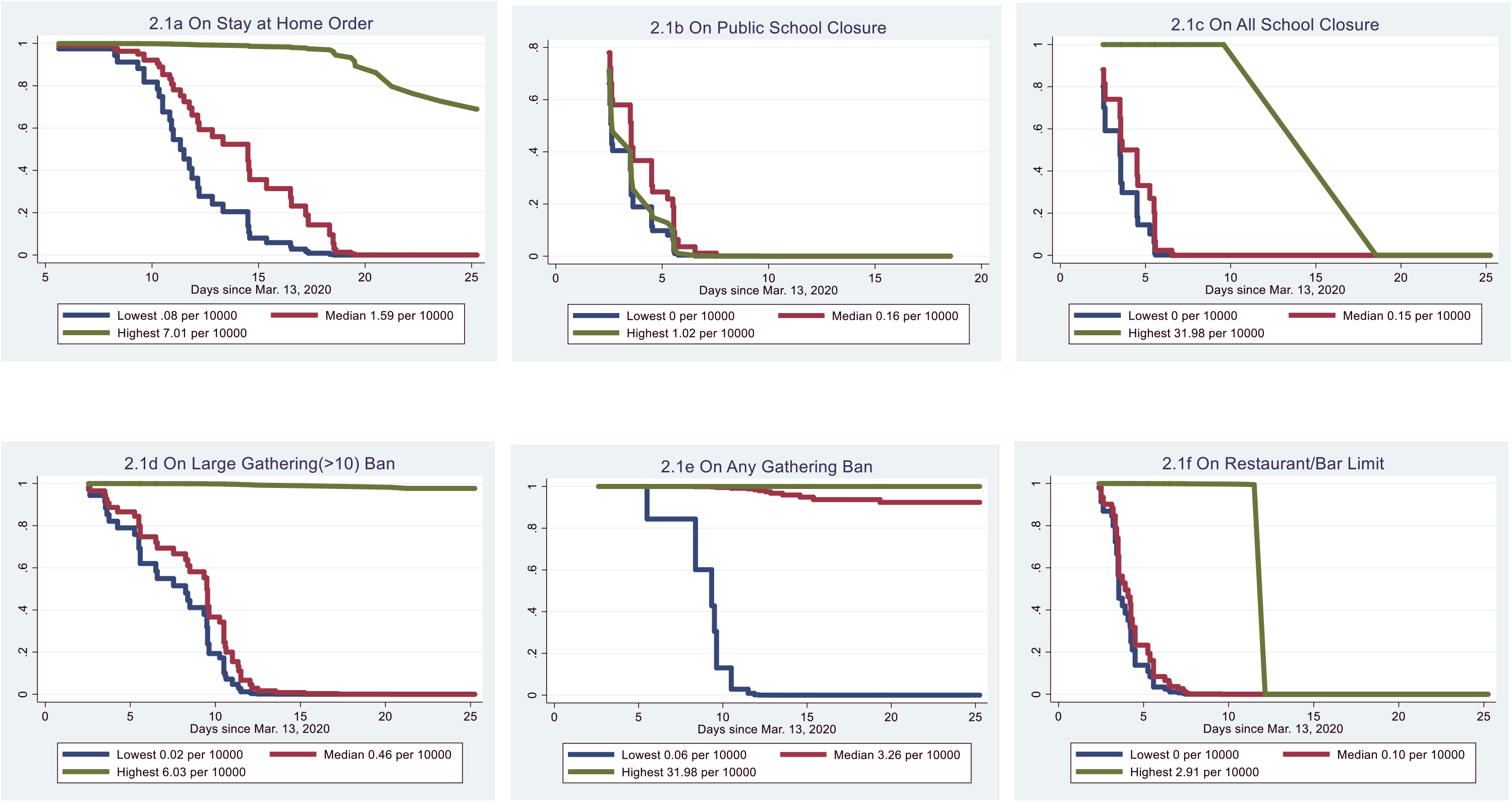
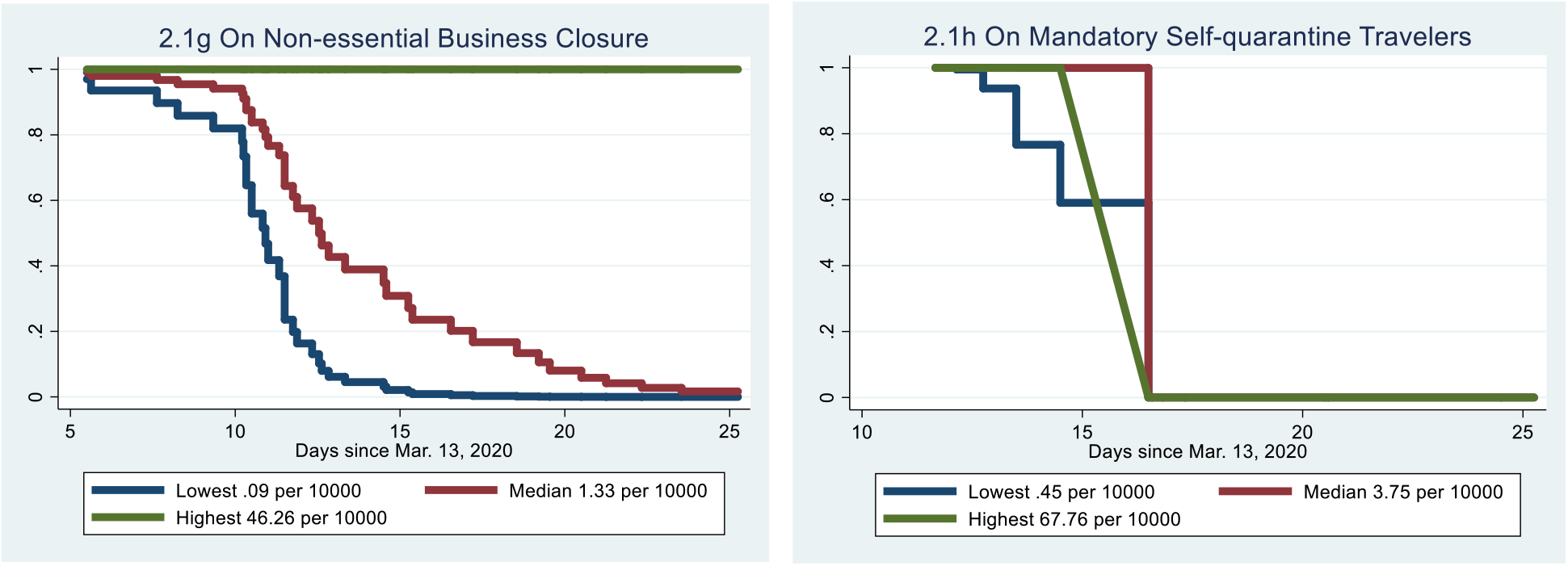
Predicted Proportions Remaining in Not Issuing Order and Restrictions by Predictors: The Impact of Covid-19 Cases per 10,000 Population on the Day prior to Enacting the Measure

**Fig. 2.2.**
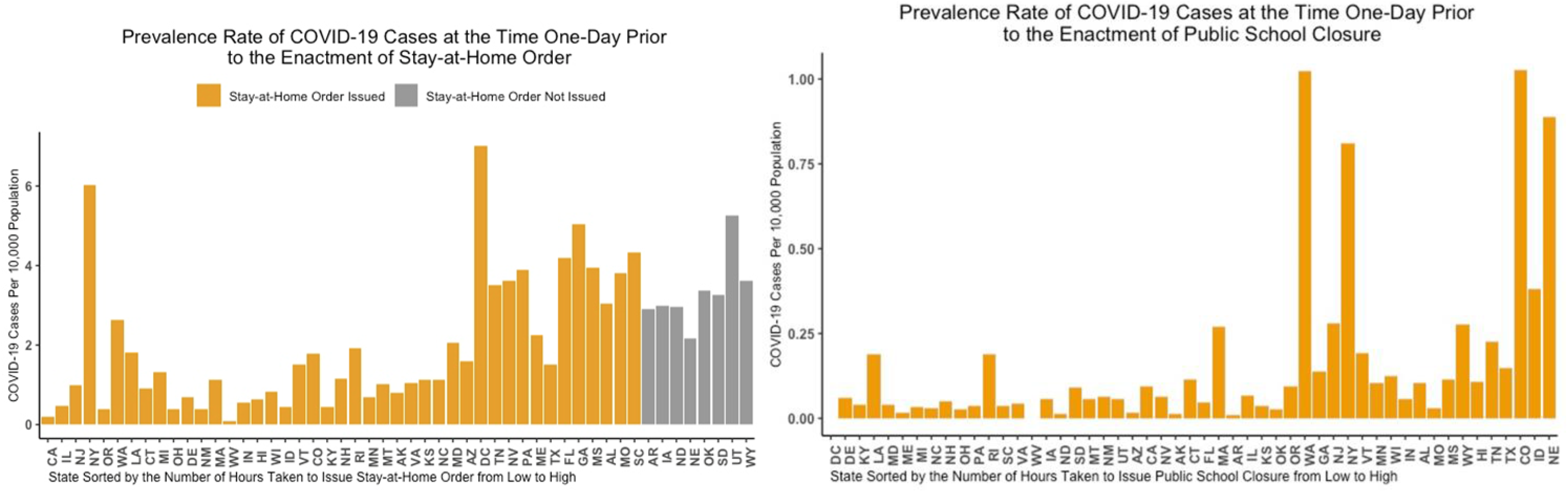
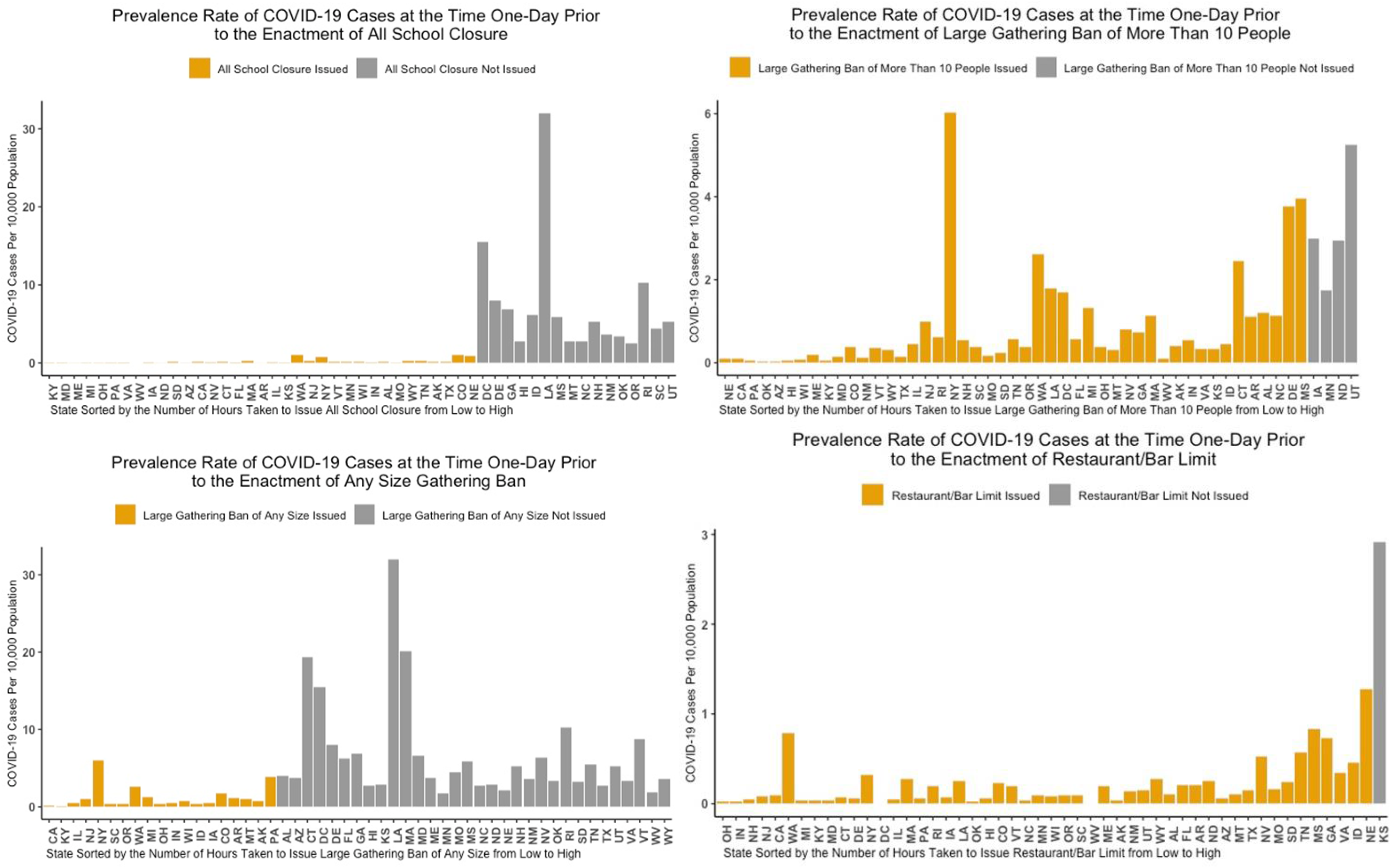
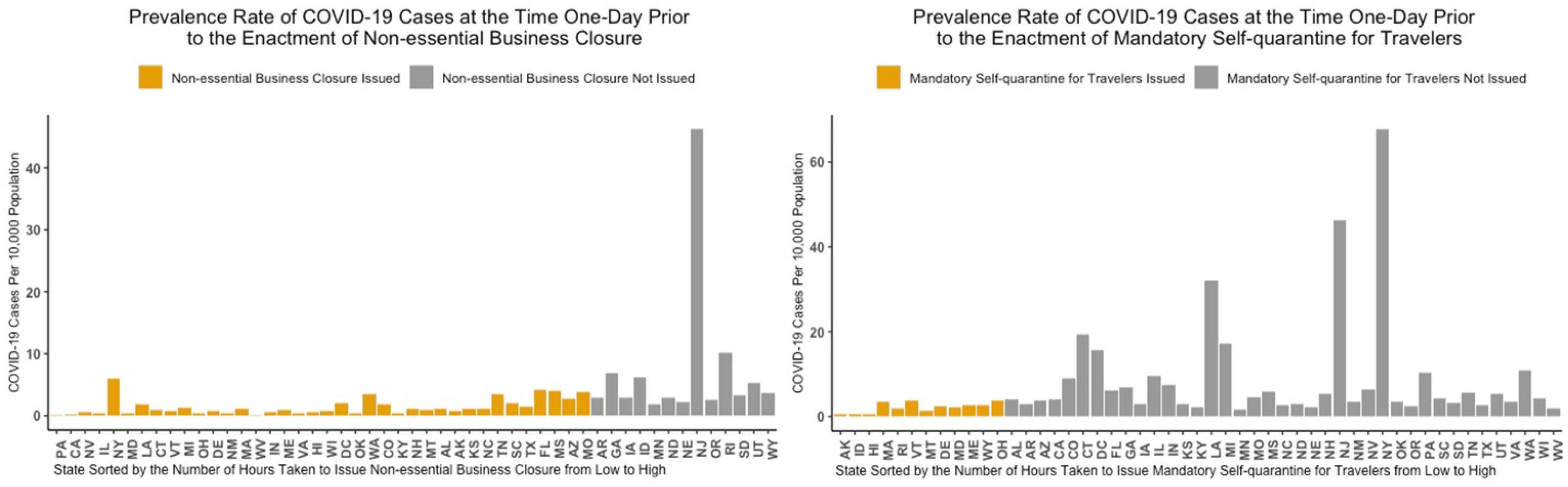
Prevalence Rates of Covid-19 Cases at the Time One Day prior to Each Mitigation Intervention was Enacted (States on the X-axis Are Arranged by Timing)

### The Impacts of the Mitigation Measures

Table 3 presents the results of the exploratory impact study. We conducted one-tailed tests on all coefficients, because based on a strong theoretical justification, we hypothesized that each independent variable (i.e., the timing of enacting a mitigating measure) reduces the new Covid-19 cases or deaths. Results show that of 72 regression coefficients (i.e., 8 mitigation measures by 9 outcome variables), only five were significantly related to mitigation strategies (p<.05, one-tailed test). All other coefficients are not statistically significant.

**Table 3.** The Impacts of Social Distancing Restrictions on the Covid-19 Cases, Deaths, New Cases, New Deaths, and Related Rates: Results from Random-effect Spatial Error Panel Model with Indirect-INT

|  | Cumulative cases |  |  | Cumulative deaths |  |  | New cases |  |  | New deaths |  |  |
| --- | --- | --- | --- | --- | --- | --- | --- | --- | --- | --- | --- | --- |
| Variables | Estimate | 95% C.I. |  | Estimate | 95% C.I. |  | Estimate | 95% C.I. |  | Estimate | 95% C.I. |  |
| Intercept | -0.003 | -0.322 | 0.316 | 0.000 | -0.236 | 0.236 | -0.002 | -0.254 | 0.251 | 0.000 | -0.206 | 0.206 |
| Stay at home order | 0.093 | 0.000 | 0.187 | 0.072 | -0.071 | 0.216 | 0.041 | -0.080 | 0.161 | 0.052 | -0.107 | 0.211 |
| Public school closure | 0.021 | -0.127 | 0.168 | -0.064 | -0.291 | 0.163 | -0.051 | -0.242 | 0.140 | -0.043 | -0.295 | 0.209 |
| All school closure | -0.050 | -0.161 | 0.060 | 0.148 | -0.023 | 0.319 | 0.047 | -0.098 | 0.192 | 0.152 | -0.039 | 0.343 |
| Large gathering ban | 0.030 | -0.053 | 0.113 | 0.049 | -0.078 | 0.175 | 0.091 | -0.015 | 0.198 | 0.025 | -0.115 | 0.165 |
| Any gathering ban | 0.036 | -0.048 | 0.120 | 0.143 | 0.018 | 0.268 | 0.174 | 0.068 | 0.279 | 0.187 | 0.051 | 0.323 |
| Restaurant/bar limit | 0.072 | -0.038 | 0.182 | 0.017 | -0.151 | 0.185 | 0.112 | -0.029 | 0.253 | 0.071 | -0.115 | 0.256 |
| Non-essential business ban | -0.234* | -0.320 | -0.149 | 0.000 | -0.132 | 0.132 | -0.137* | -0.248 | -0.026 | -0.028 | -0.174 | 0.118 |
| Mandatory self-quarantine of travelers | -0.006 | -0.113 | 0.102 | 0.148 | -0.016 | 0.311 | 0.065 | -0.072 | 0.202 | 0.094 | -0.086 | 0.274 |
| Coefficient of spatial error autocorrelation | $\rho = 0.223$, p-values = 0.000 | | | $\rho = 0.034$, p-values = 0.049 | | | $\rho = 0.076$, p-values = 0.019 | | | $\rho = -0.020$, p-values = 0.589 | | |

\* p<.05, a one-tailed test

Table 3. Continued
| Variables | Proportion of cumulative cases over the population |  |  | Proportion of Cumulative deaths over the population |  |  | Proportion of new cases over the population |  |  |
| --- | --- | --- | --- | --- | --- | --- | --- | --- | --- |
|  | Estimate | 95% C.I. |  | Estimate | 95% C.I. |  | Estimate | 95% C.I. |  |
| Intercept | -0.004 | -0.293 | 0.284 | -0.001 | -0.241 | 0.240 | 0.000 | -0.205 | 0.205 |
| Stay at home order | 0.170 | 0.054 | 0.286 | 0.046 | -0.102 | 0.194 | 0.143 | -0.023 | 0.308 |
| Public school closure | -0.080 | -0.263 | 0.103 | -0.002 | -0.236 | 0.232 | -0.137 | -0.399 | 0.125 |
| All school closure | 0.116 | -0.020 | 0.252 | 0.204 | 0.029 | 0.379 | 0.223 | 0.026 | 0.420 |
| Large gathering ban | 0.023 | -0.080 | 0.126 | 0.027 | -0.105 | 0.158 | 0.161 | 0.015 | 0.307 |
| Any gathering ban | 0.035 | -0.069 | 0.139 | 0.087 | -0.043 | 0.218 | 0.070 | -0.073 | 0.212 |
| Restaurant/bar limit | 0.143 | 0.007 | 0.279 | -0.055 | -0.229 | 0.119 | 0.120 | -0.074 | 0.313 |
| Non-essential business ban | -0.138* | -0.244 | -0.031 | 0.047 | -0.089 | 0.183 | -0.092 | -0.244 | 0.060 |
| Mandatory self-quarantine of travelers | 0.133 | 0.000 | 0.266 | 0.363 | 0.193 | 0.532 | 0.256 | 0.068 | 0.444 |
| Coefficient of spatial error autocorrelation | $\rho = 0.214$, p-values = 0.000 | | | $\rho = 0.116$, p-values = 0.000 | | | $\rho = 0.015$, p-values = 0.651 | | |

\* $p < .05$ , a one-tailed test

Table 3. Continued
|  | Proportion of new deaths<br>over the population |  |  | Death rate |  |  |
| --- | --- | --- | --- | --- | --- | --- |
| Variables | Estimate | 95% C.I. |  | Estimate | 95% C.I. |  |
| Intercept | 0.000 | -0.187 | 0.187 | -0.024 | -0.193 | 0.145 |
| Stay at home order | 0.093 | -0.093 | 0.278 | 0.037 | -0.130 | 0.203 |
| Public school closure | -0.078 | -0.370 | 0.213 | -0.098 | -0.363 | 0.166 |
| All school closure | 0.183 | -0.034 | 0.400 | 0.452 | 0.252 | 0.652 |
| Large gathering ban | 0.013 | -0.151 | 0.177 | -0.374* | -0.522 | -0.227 |
| Any gathering ban | 0.141 | -0.018 | 0.300 | 0.338 | 0.197 | 0.479 |
| Restaurant/bar limit | -0.026 | -0.243 | 0.191 | -0.234* | -0.429 | -0.039 |
| Non-essential business ban | -0.029 | -0.200 | 0.141 | 0.460 | 0.306 | 0.614 |
| Mandatory self-quarantine<br>of travelers | 0.299 | 0.089 | 0.510 | 0.312 | 0.123 | 0.501 |
| Coefficient of spatial error<br>autocorrelation | $\rho = 0.011$, p-values =<br>0.656 | | | $\rho = -0.077$, p-values =<br>0.024 | | |

\* $p < .05$ , one-tailed test

The following five coefficients suggest that mitigation interventions are effective in reducing Covid-19 case data. That is, other things being equal, enacting a non-essential business ban reduces 0.228 daily cumulative cases, reduces 0.133 daily new cases, and reduces 0.134 daily cumulative cases per 10,000 population; enacting a large-gathering ban reduces the daily Covid-19 death rate by 35.5%; and enacting a restaurant/bar limit reduces the daily Covid-19 death rate by 22.3%.

In summary, three mitigation strategies (i.e., non-essential business ban, large-gathering ban of more than 10 people, and restaurant/bar limit) may be effective in reducing cumulative cases, new cases, and death rate. All three strategies aim to increase social distance by reducing social gatherings and contact. Other social distancing strategies were not correlated with outcomes, and it is unknown whether effects would be observed over a greater intervention period.

## Discussion

To date, the United States continues to experience high levels of Covid-19 disease. This study offers two observations that may help explain why this is the case. One is that some states may have missed optimal timing to implement social distancing. Based on prior research during epidemics, it is important to implement social distancing swiftly; the key is not whether such measures should be taken but when. The second is that, in the short run of 5 weeks, mitigation interventions should be expected to have only limited effects on slowing down the spread of disease. Of nine strategies, only three showed impacts.

Four implications of these findings warrant special attention. First, Stay-at-Home orders, public school closures, all school closures, any gathering bans, mandatary self-quarantine of travelers are important, but perhaps – in the early stages of trying to control a highly infectious disease – are not as important as the large-gathering ban of more than 10 people, non-essential business closure, and restaurant/bar limits. These three restrictions focus on avoiding large gatherings and interacting crowds. Observing an effect from other mitigation strategies may require more time. School closures, for example, may take a longer time to demonstrate an effect because, in this epidemic, young people appear to have been less affected, and they were more likely to be asymptomatic.

Second, staying at home may not reduce new disease if home is a collective, such as a long term care facility, and if exposure occurs in home settings. In this regard, very quick tracing may be necessary. China’s public-private “health code” system may be worthy of consideration. Hosted on the Alipay and WeChat apps and deeply aligned with the public health system, a health code of one of three viral risk statuses – red, yellow, or green – is assigned to every citizen.^19^ This enables quicker tracking and quarantining than is possible – and perhaps legally feasible – in the Western countries, where privacy concerns often dominate discourse on public health interventions. Notwithstanding, applying advanced technology, using big data, and mobilizing digital tools to better target health care system resources is recommended by the National Academies.^9^

Thirdly, during the study window, many U.S. policymakers and officials considered reopening the economy by removing some or all mitigation interventions. As of May 11, or about three weeks after the study window was closed, 32 states did so.^6^ Our exploratory findings suggest that at least three mitigation strategies were having an effect at that time. Because there is no preventive vaccine and because there are few potentially effective treatments, recent reductions in new cases and deaths must be due, in large part, to the social interventions delivered by states. The removal of these social interventions in the absence of effective pharmaceutical interventions carries a high risk for a relapse.

Finally, it is worth noting that this analysis is based on the decisions made by Governors at the state level so may imprecisely measure the exact variation of impacts across the U.S., since in many cases social distancing policies were implemented at the substate level (e.g. counties) at different periods, reflecting different mitigation strategies and leading to different outcomes.

The unprecedented implementation of social distancing is projected to cost at least 2 trillion U.S. dollars. On March 27, an over $2 trillion economic relief package, the Coronavirus Aid, Relief, and Economic Security (CARES) Act, was passed by Congress with bipartisan support, and it was signed into law by President Trump.^20^ Social distancing interventions are also associated with a large number of job losses. In the seven weeks ending May 2, 33.5 million people made initial claims for unemployment in the U.S., a record number of unemployment claims. The official unemployment rate reached 14.7% in April, the highest rates observed since the Great Depression.^21^ Undoubtedly, policymakers are facing a tough decision: How to balance between economic and public health interests? Which strategies should be undertaken at which time points in order to maximize utility functions? The findings of this study provide preliminary evidence about the effectiveness of some mitigation measures.

Policymakers need to weigh carefully the pros and cons of lifting the mitigations, given that these measures were already associated with a huge cost and economic resources.

## Data Availability

All data employed in this study are from governmental websites, census data, and the Johns Hopkins University Coronavirus Data Stream that combines WHO and CDC case data

## Acknowledgments

We thank Mary M. McKay for her leadership in transdisciplinary research integrating public health, social work, and social policy. We thank Mark W. Fraser, Danyu Lin, Zachary R. McCaw for their excellent suggestions and comments on this study, and Peter Sun for his assistance with data management.

## Supplementary Materials

### The Correction of a Non-normal Outcome Variable with Inverse-Normal Transformation

Results show that the inverse-normal transformation method appropriately transformed the zero-inflated and skewed outcome into a normally distributed variable. Fig. S1 shows an example.

**Figure S1.**
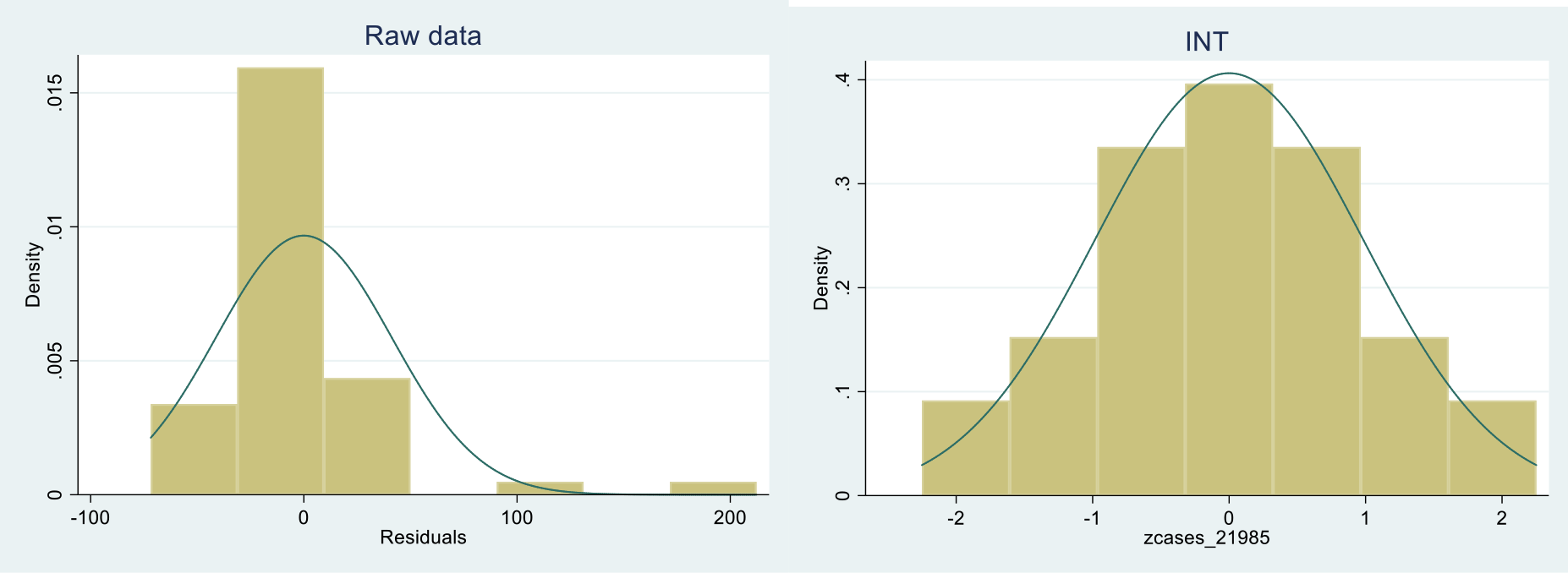
Distribution of an outcome variable before and after INT using the Covid-19 Cases on March 13, 2020 as an example

### Additional Model-based Predictions of Survival Function

Figure 2 presents the model-based predictions of survival curve showing the effect of prevalence rate on the timing of enacting mitigation interventions. Since the results of other significant predictors are also important, particularly for policymakers and stakeholders, we present selected model-based predictions of survival curves in Figure S2. In these figures (S2.1-S2.6), the bottom survival line indicates the fastest speed to enact, while the top survival line indicates the slowest.

**Figure S2.**
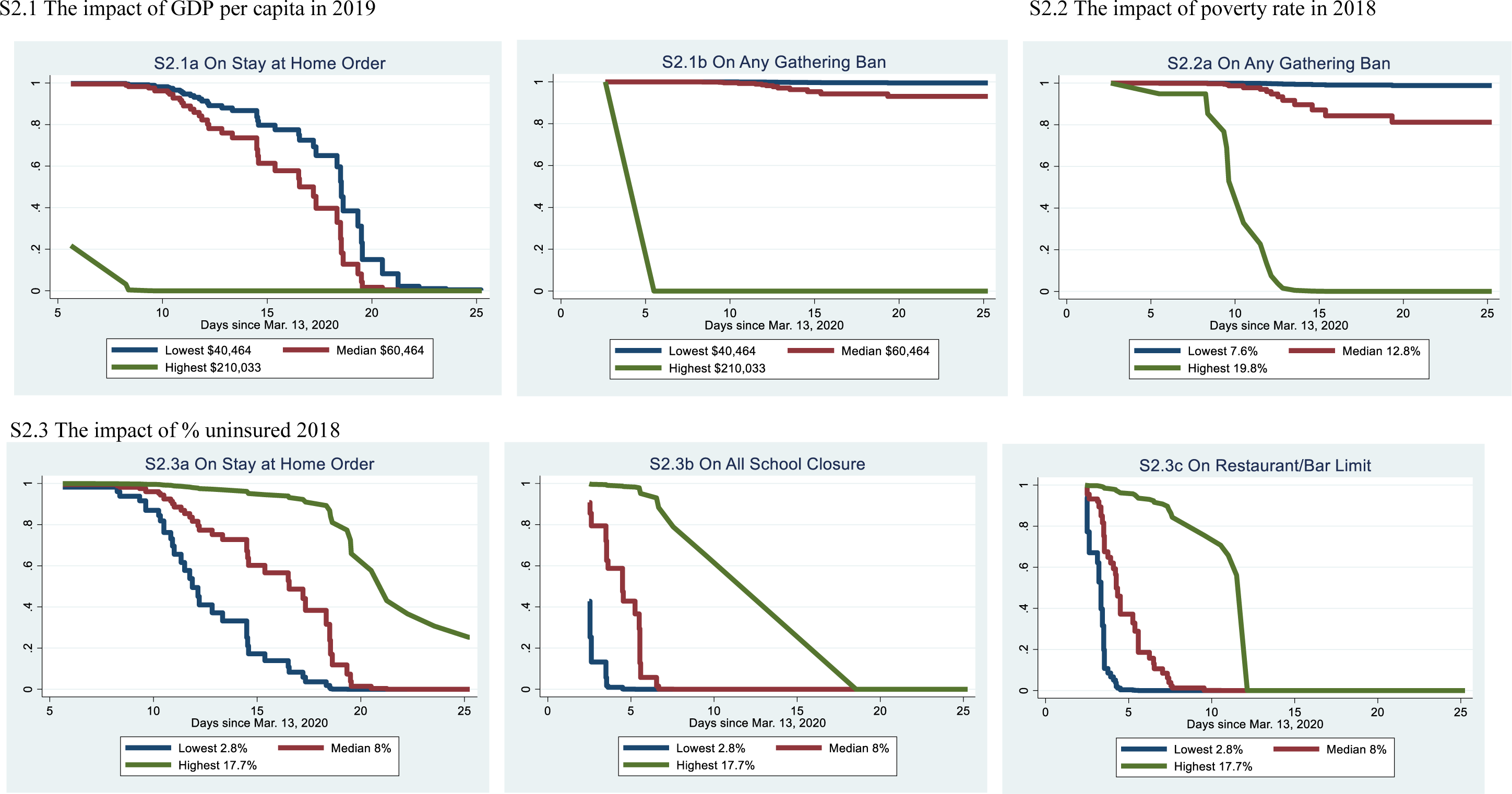
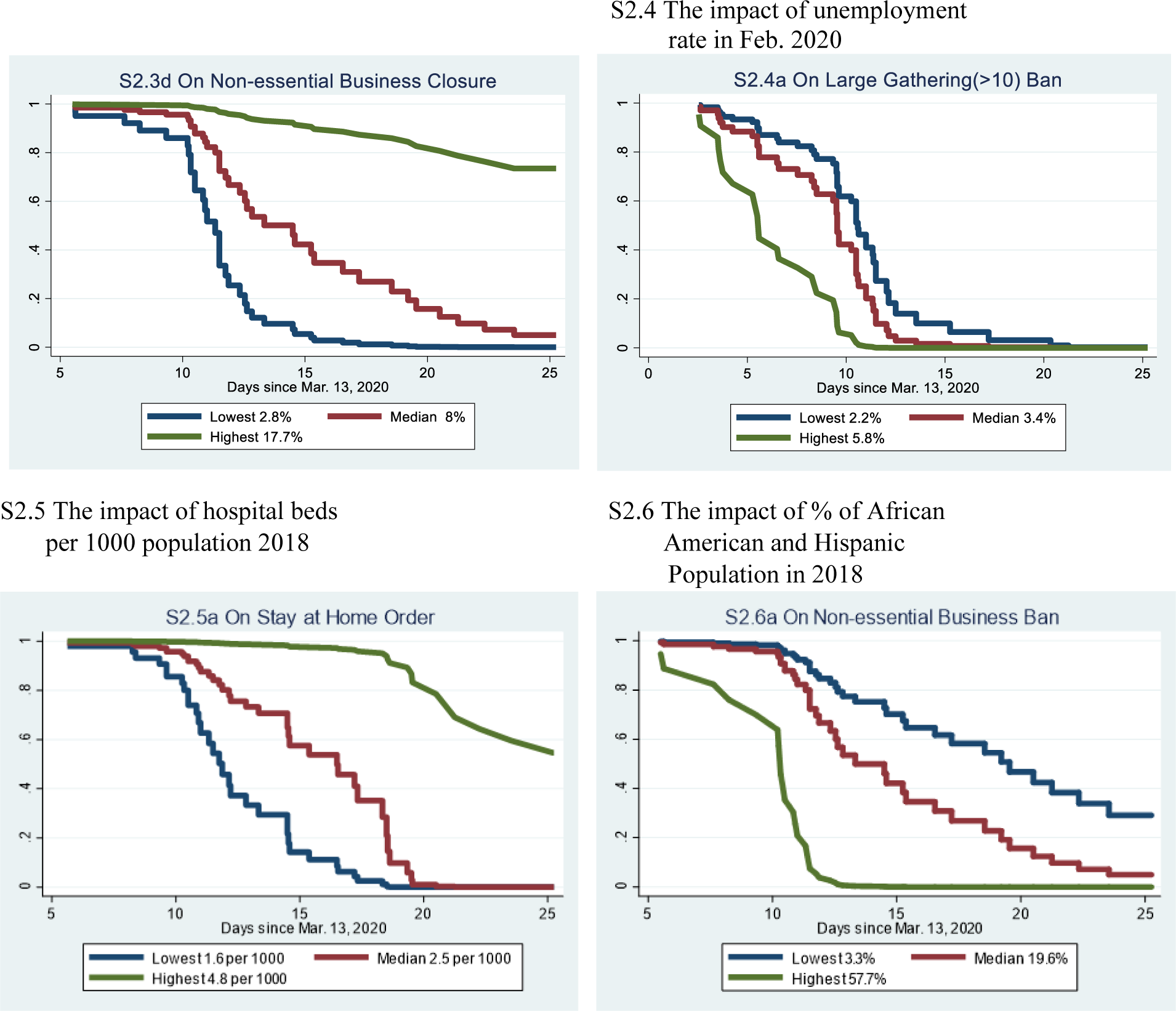
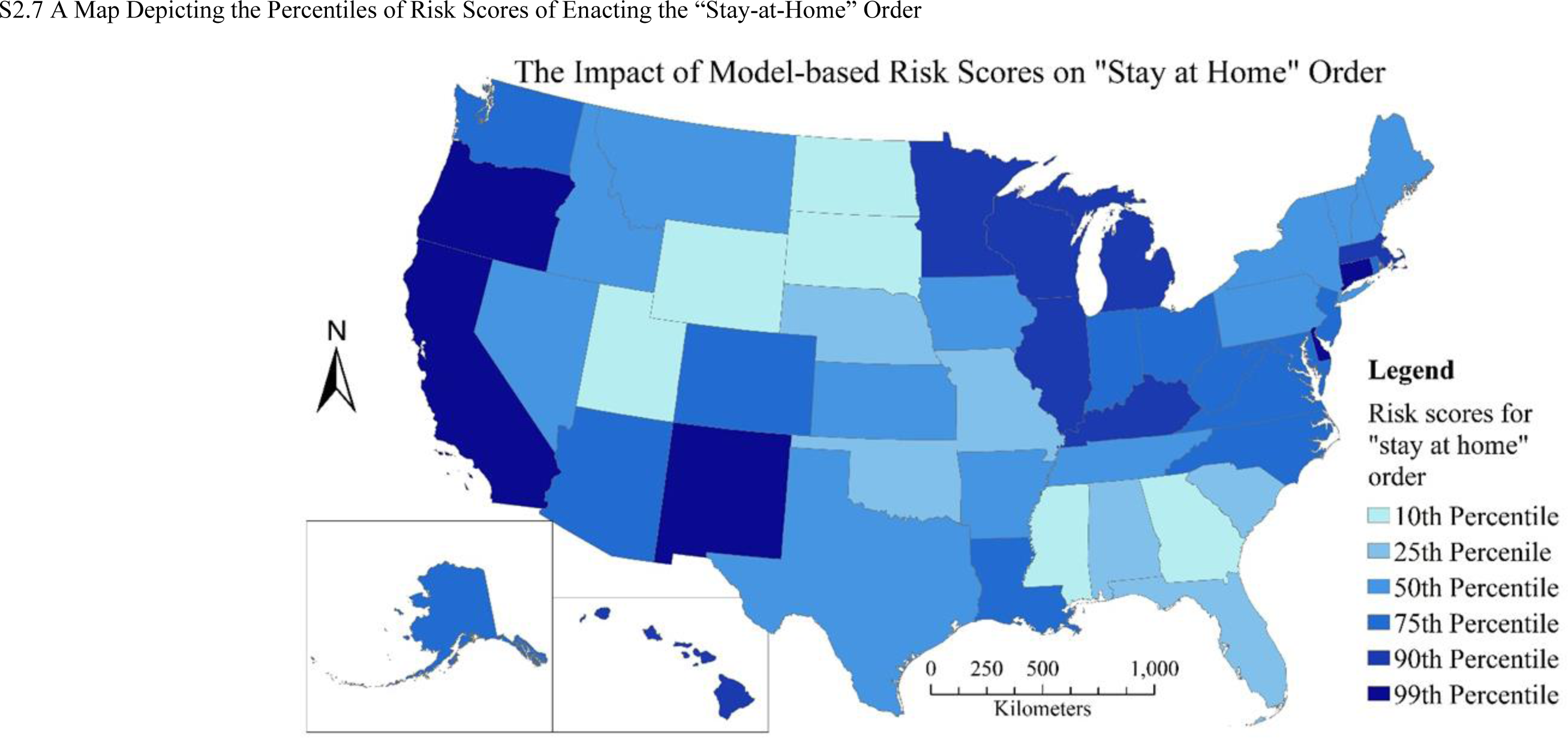
Predicted Proportions Remaining in Not Issuing Order and Restrictions by Selected Predictors

Figure S2.7 shows a map of model-based prediction of the percentiles of risk scores issuing the Stay-at-Home order. These scores were obtained by multiplying the values of independent variables for each of the 50 states and Washington DC by the estimated regression coefficients of the proportional hazards model. Risk score in the current setting indicates the speed enacting the Stay-at-Home order, with states of the 10th percentile moving to enact the order the slowest speed or having the highest proportion not enacting the order at a given time, and states of the 99th percentile moving to enacting the order the fastest speed.

### Sensitivity Analysis of the Impact of Prevalence Rate on Mitigation Timing

Findings of the predictor of Covid-19 cases per 10,000 population, or the prevalence rate, are contradictory to our hypothesis: states with a lower prevalence of Covid-19 cases per 10,000 population reacted more quickly to the outbreak. To ensure that this pattern did exist, we conducted several sensitivity analyses. We first replaced this variable with the growth rate of Covid-19 cases per 10,000 population from five days ago to one day ago, in conjunction with the case count five days ago. We tested both the linear growth rate and exponential grown rate. The analysis was performed on all eight mitigation measures. Results from these analyses confirmed that states with high prevalence rates enacted mitigation strategies slowly.

### A Note about Nonsignificant Impacts of the Stay-at-Home Order

Surprisingly, the results from Table 3 seem to suggest that the “Stay-at-Home” order does not have significant impact on any of the nine outcome variables. We suspect that the “Stay-at-Home” order does not necessarily confine people from contacting one another since the order is more like the opposite of “go-to-work.” The “Stay-at-Home” order hence might not be a good measure for “social distancing” as was intended. While the order was issued, most will simply not go to work, but gather as they would do after work. The three significant factors that are found in the four models, non-essential business closure, large-gathering ban, restaurant/bar limit, are probably the real measures for “social distancing.” We experimented with the same random effect spatial error panel models but removed the three significant predictors. The modified models still suggest that the “Stay-at-Home” order does not have a significant impact on any of the outcome variables. These results suggest that “social distancing” strategy indeed works against the spread of Covid19. While “stay-at-home” order was initially thought as a means for social distancing, it fails to achieve real social distancing nationwide.

In addition, the timing of when the “Stay-at-Home” order was issued also suggests a spatial clustering pattern (Figure 1). A spatial autocorrelation test of the timing when the “Stay-at-Home” order was issued produces a Moran’s I of 0.348, with a p-value = 0.00024 using the SOI spatial configuration. The test suggests that the timing of issuing “Stay-at-Home” order might have a clear spatial clustering trend in which one state is more likely to enact the “Stay-at-Home” order if its neighbors are enacting the order and vice versa.

